# A two-phase stochastic dynamic model for COVID-19 mid-term policy recommendations in Greece: a pathway towards mass vaccination

**DOI:** 10.1101/2021.01.07.21249394

**Authors:** N.P. Rachaniotis, T.K. Dasaklis, F. Fotopoulos, P. Tinios

## Abstract

From November 7^th^, 2020, Greece adopted a second nationwide lockdown policy to mitigate the transmission of SARS-CoV-2 (the first took place from March 23^rd^ till May 4^th^, 2020), just as the second wave of COVID-19 was advancing, as did other European countries. In the light of the very promising voluntary mass vaccination, which will start in January 2021, it is of utmost importance for the country to plan to complement vaccination with mid-term Non-Pharmaceutical Interventions (NPIs). The objective is to minimize human losses and to limit social and economic costs. In this paper a two-phase stochastic dynamic network compartmental model (a pre-vaccination SEIR until February 15^th^, 2021 and a post-vaccination SVEIR from February 15^th^, 2021 to June 30^th^, 2021) is developed. Three scenarios are assessed in the first phase: (a) a *baseline scenario*, which lifts the national lockdown and all NPIs on January 2021, (b) a *“semi-lockdown*” scenario with school opening, partial retail sector operation, universal mask wearing and social distancing/teleworking on January 2021 and (c) a “*rolling lockdown*” scenario combining a *partial lifting of measures* in January 2021 followed by a third imposed nationwide lockdown in February 2021. In the second phase three scenarios with different vaccination rates are assessed. Publicly available data along with some preliminary first results of the SHARE COVID-19 survey conducted in Greece are used as input. The results regarding the first phase indicate that the “semi-lockdown” scenario outperforms the third lockdown scenario (5.7% less expected fatalities), whereas in the second phase it is of great importance to ensure a sufficient vaccine supply and high vaccination rates.

## 1. Introduction

The global spread of SARS-CoV-2, the pathogen that causes the disease COVID-19, has created an infectious disease crisis worldwide straining healthcare systems to their limits. The COVID-19 outbreak presents the greatest challenge after World War II. As its repercussions quickly spilled over from healthcare into other sectors such as global logistics and trade, travel and tourism, retail, energy and finance, it has wreaked havoc in health, social and economic systems. Until December 31st, 2020, at a global scale, 81,947,503 confirmed cases of COVID-19, including 1,808,041 deaths^1^ were reported. In Greece, from the onset of the outbreak (February 2020) until December 31^st^, 2020, 138,850 confirmed cases of COVID-19 were reported respectively, including 4,838 deaths^2^.

In the absence of specific medical anti-COVID-19 treatment protocols until December 2020, the vast majority of countries around the globe adopted various Non-Pharmaceutical Interventions (NPIs) to control the spread of the disease and to minimize death rates. NPIs adopted in Greece have included so far: social distancing and lockdowns, travel restrictions, closure of schools and universities, avoidance of mass gatherings, point of entry screening, teleworking, community-wide containment, iterative contact tracing, quarantining and self-isolation strategies, etc. Several promising vaccine candidates are under development (many in phase 3 of clinical trials) and some have already been licensed. As of December 31^st^, 2020, the following six types of vaccine candidates have been developed and are currently under clinical trials: non-replicating viral vectors (NRVV), messenger RNA vaccine candidates, self-amplifying messenger RNA vaccine candidates, DNA vaccine candidates, inactivated whole-virus vaccine candidates and protein subunit vaccine candidates [1]. Moreover, in December 31, 2020, the World Health Organization (WHO) listed the Comirnaty COVID-19 mRNA vaccine for emergency use, making the Pfizer/BioNTech vaccine the first to receive emergency validation from WHO since the beginning of the COVID-19 outbreak.

As vaccines are gradually becoming available, countries worldwide have started developing exit strategies from the COVID-19 outbreak. Such strategies intend to strike a balance between the use of limited vaccine resources (as they become gradually available and subject to logistical and distribution constraints) and the utilisation of various NPIs, while at the same time minimizing mortality rates and the negative effects of prolonged NPIs to the society and the economy. Several mathematical epidemiological models have been proposed in the literature for capturing COVID-19 disease dynamics and assess control policies. These are categorized in two broad classes: deterministic and stochastic. Typical deterministic compartmental models are usually based on a set of differential equations describing transition dynamics between different compartments, assuming uniform (homogeneous) mixing of the population. A deterministic model will always yield the same output from a given initial condition. On the other hand, stochastic models incorporate the inherent stochasticity of infectious disease outbreaks and also account for heterogeneous mixing patterns of the affected population. Note that it remains crucial to consider stochasticity, heterogeneity and the structure of the population contact networks when studying COVID-19 transmission, given that containment strategies are typically implemented through perturbing these networks (e.g. social distancing) or exploiting them (e.g. contact tracing). Therefore, stochastic models could be extensively used to assess the utility of specific NPIs or the accrued benefits of combining NPIs and pharmaceutical interventions (such as usage of vaccines).

In this paper a two-phase stochastic dynamic network compartmental model (a pre-vaccination Susceptible-Exposed-Infected-Recovered (SEIR) until February 15^th^, 2021 and a post-vaccination Susceptible-Vaccinated-Exposed-Infected-Recovered (SVEIR) from February 15^th^, 2021 to June 30^th^, 2021) is developed, implemented and tested. *Three* scenarios are assessed in the first phase: (a) a baseline scenario lifting the national lockdown and all NPIs at January 8^th^, 2021, (b) a “semi-lockdown” scenario with school opening in January 8^th^, 2021, partial retail sector operating, universal mask wearing and social distancing/teleworking and, (c) a “*rolling lockdown*” scenario combining a partial lifting of measures on January 8^th^ with a third imposed nationwide lockdown in February 2021. In the *second* phase three scenarios with different vaccination rates are assessed. Publicly available data along with the preliminary results of the SHARE COVID-19 survey in Greece are used as input. The results are showcasing mid-term policy recommendations regarding the pandemic containment in Greece and could support decision makers in suggesting a pathway until the majority of the population is vaccinated.

The remainder of this paper is organized as follows. Section 2 provides a comprehensive review of the available stochastic COVID-19 epidemiological models with a specific focus on studies incorporating vaccination-related strategies. In Section 3, the proposed two-phase stochastic model is described, whereas in Section 4, the experimental results of the simulation runs performed are presented. Finally, in Section 5, the various benefits of the proposed model along with the relevant policy-oriented implications of the obtained results are discussed.

## 2. Background literature

Starting from the first days of 2020, COVID19-related epidemiological literature is abundant. Various models have been proposed so far in order to capture its transmission mechanism and characteristics [2], [3]. However, given that an effective vaccine was not available until recently, most studies have primarily focused on modeling NPIs for controlling the COVID19 outbreak. Such interventions include quarantine over certain regions, social distancing, leisure closure, masking, etc. Of particular interest are stochastic epidemic models that, in contrast to deterministic models, capture disease transmission characteristics along with the heterogeneity of population dynamics. Stochasticity in disease dynamics can be captured either by agent-based models [4], [5] or by stochastic compartmental models [6], [7].

The advent of COVID19-related vaccines has a significant impact on modelling and assessing relevant control policies. Two main streams of vaccine-related papers within the COVID19 literature have been identified: The first stream addresses issues of vaccine prioritization (across different age groups, regions, timeframes, etc.), minimum immunization thresholds for achieving herd immunity and type of immunization campaign undertaken (impulsive/voluntary vaccination, etc.). For example, in [8] the authors assess the optimal allocation of a limited vaccine supply across different age groups. The authors investigate three alternative policy objectives (minimizing infections, years of life lost, or deaths) and they further demonstrate how optimal prioritization is responsive to many parameters, most notably the efficacy and availability of vaccines, the rate of transmission and the severity of initial infections. An information-dependent model for assessing behavioral aspects of an immunization campaign is presented in [9]. The overall vaccination process is presumed to be completely voluntary and it is assumed that the decision to get vaccinated or not depends in part on the knowledge and rumors available about the spread of the disease in the population. Other studies identify the necessary vaccine stockpile level for achieving herd immunity within a given population for different levels of vaccine’s efficiency [10][11]. In [12] the authors suggest a time- and space-based vaccine distribution strategy that sequentially prioritizes regions with the most recent cases of infection within a certain time period and contrasts it with the current practice of demographically distributing vaccines. The suggested vaccine distribution plan sets out the idea that individuals should be prioritized not only by individual characteristics, such as their risk of spreading the disease, but also by the area in which they reside. Two optimal control problems (single- and multi-objective) are suggested to assess a vaccine administration strategy for COVID-19 in [13]. The first strategy is to minimize the number of people infected during the course of treatment whereas the second considers that the sum of affected individuals and the prescribed vaccine concentration should be reduced together during the control effort. Finally, in [14] the authors assess two vaccination strategies i.e. newborns vaccination and voluntary vaccination.

The second stream focuses on the possible benefits of combining vaccination with NPIs such as surveillance, social distancing, social relaxation, quarantining, patient treatment/isolation, etc. for various levels of vaccine efficiency [15]. In [16] the authors construct a game-theoretic model for disease transmission and test two control measures (i.e. vaccination and social distancing). In [17] the authors examine the combination of immunization campaigns with other control measures such as masking and social distancing. Vaccination may prove critical when considering disease dynamics within subpopulations that are extremely vulnerable to COVID19 (such as the elderly or institutionalized individuals). Such groups may become hotspots driving infection within the general population [18]. Targeted vaccination of these groups may be sufficient to interrupt regional spread and protect a much wider fraction of the public. In [19] the authors identify the necessary number of vaccines and vaccine efficacy thresholds capable of preventing an epidemic whilst adhering to lockdown guidelines. Assuming a vaccine efficacy of 100% in a mass vaccination program, at least 60% of a given population should be vaccinated to obtain herd immunity. It should be noted that for eliminating disease transmission a highly effective vaccine is required and, therefore, combining vaccination with other interventions, such as face mask usage and/or social distancing always yield optimal containment results [20].

The proposed two-phase (a pre-vaccination SEIR and a post-vaccination SVEIR) stochastic dynamic network based compartmental model is used to forecast the relative impact of a set of major NPIs, which can be specified on a targeted individual basis, along with voluntary mass vaccination. The network model enables rigorous analysis of transmission patterns and network-based interventions with respect to the properties of realistic contact networks.

## 3. The model

### Phase-1 model

A modified version of the extended Susceptible-Exposed-Infectious-Recovered (SEIR) model^3^ is used to represent SARS-CoV-2 disease states (Figure 1) in the first phase where pharmaceutical interventions (vaccines) are not available. Susceptible individuals (*S*) from a population of size *N* become exposed (*E*) if they make a transmissive contact with already infectious individuals. These newly exposed individuals first experience a latent period during which they are not infectious. Then they progress to a pre-symptomatic infectious state (*I*_*pre*_), where they are infectious but asymptomatic. A percentage develops symptoms (*I*_*sym*_), while the remainder never develops symptoms despite being infectious (*I*_*asym*_). A subset of symptomatic individuals progress to a more severe clinical state and must be hospitalized (*I*_*H*_), and a fraction of these severe cases are fatal (*F*). At the end of the infectious period, infected individuals enter the recovered state (*R*) and are no longer infectious or susceptible to infection. It must be stated here that recovered individuals may become resusceptible sometime after recovering, though with a highly uncertain and in any case very low rate [21]. Transmissibility, rates of progression and other properties vary between the disease states.

**Figure 1.**
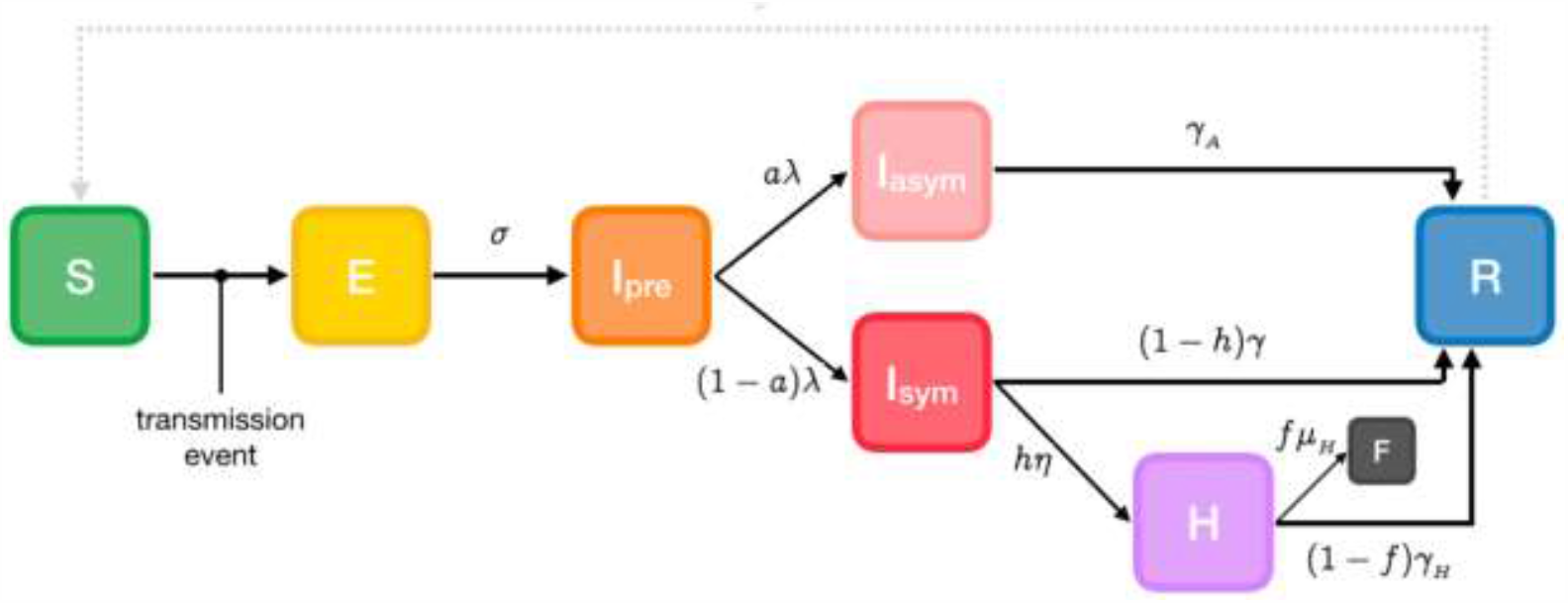
Phase-1 SEIR model^4^.

### Parameters

The model’s parameters are the following:

- *α*: susceptibility
- *β*: transmissibility of symptomatic individuals
- *β*_*A*_: transmissibility of pre- and asymptomatic individuals
- *σ*: rate of progression to infectiousness (1/latent period)
- *λ*: rate of progression to (a)symptomatic state (1/pre-symptomatic period)
- *a*: probability of an infected individual remaining asymptomatic
- *h*: probability of a symptomatic individual being hospitalized
- *η*: rate of progression to hospitalized state (1/onset-to-admission period)
- *γ*: rate of recovery for non-hospitalized symptomatic individuals (1/symptomatic individuals infectious period)
- *γ*_*A*_: rate of recovery for asymptomatic individuals (1/asymptomatic individuals infectious period)
- *γ*_*H*_: rate of recovery for hospitalized symptomatic individuals (1/hospitalized individuals infectious period)
- *f*: probability of death for hospitalized individuals (case fatality rate)
- *μ*_*H*_: rate of death for hospitalized individuals (1/hospital admission to death period)
- *ξ*: rate of re-susceptibility (1/temporary immunity period; equal to 0 if a permanent immunity period is assumed)

### Phase-2 model

Worldwide vaccination, which started at the end of December 2020, is planned to take place through hospitals, health centers and other primary health care infrastructure. Moreover, mobile medical teams will vaccinate people unable to proceed to vaccination centers either because they are house-bound (elderly, incapacitated etc.) or they are in institutions. To illustrate this second phase of the pandemic, an extended Susceptible-Vaccinated-Exposed-Infected-Recovered (SVEIR) model is implemented (Figure 2).

**Figure 2.**
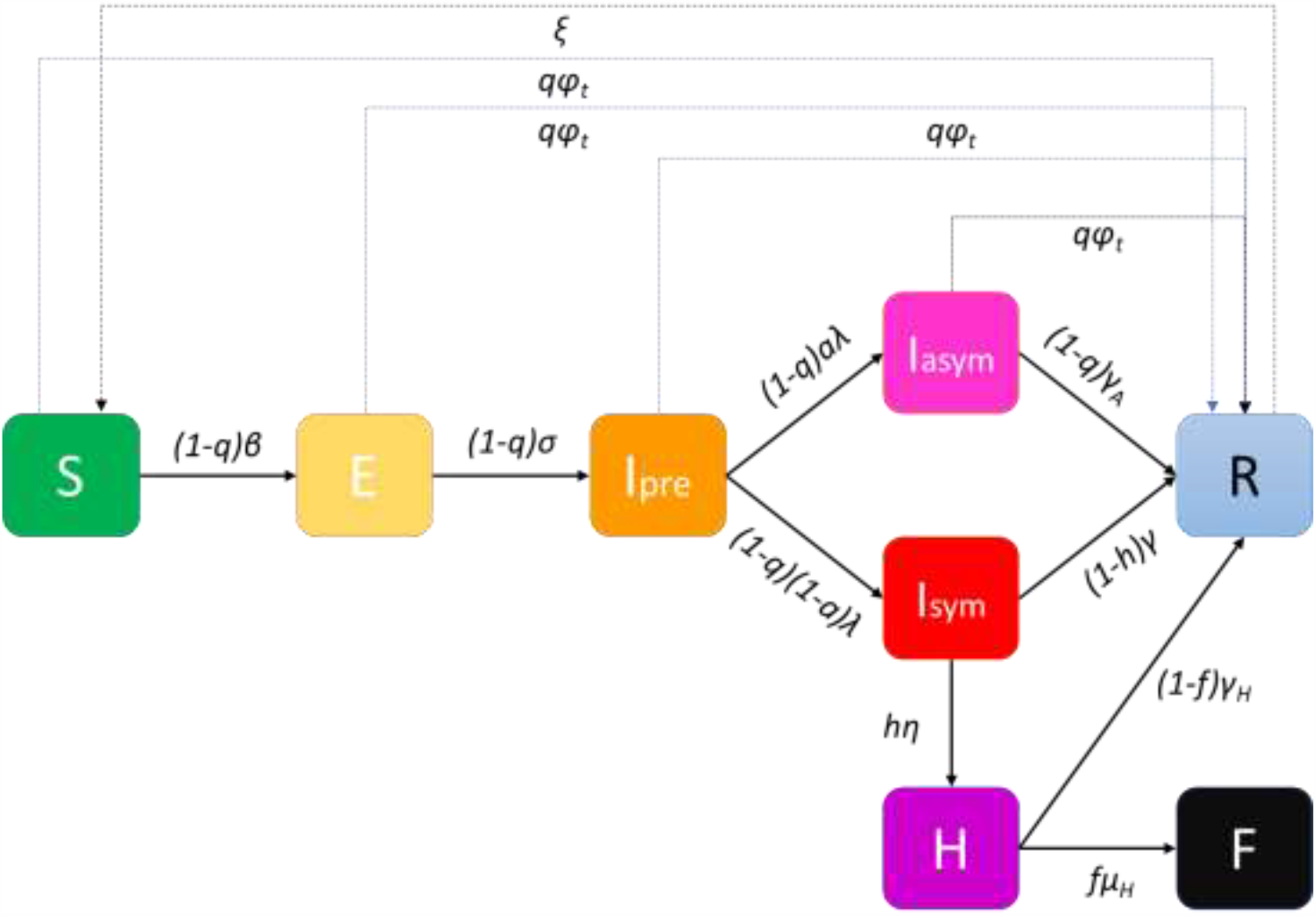
Phase-2 SVEIR model.

The number of vaccines available at all vaccination centres at different discrete time periods (usually days) *t*, a parameter that will affect the course of the outbreak, is expressed via the vaccination rate *φ*_*t*_. Vaccination will be set on a voluntary basis, and the vaccine will have an expected efficacy. This is captured in the model by a percentage *q* of the population that will be willing to participate and at the same time the vaccine will have the desired effects. As all asymptomatic individuals may present themselves to the vaccination queue, the available vaccines might be allocated to those susceptible to the disease that are eligible for vaccination and to those that are in states *I*_*pre*_ and *I*_*asym*_.

### Stochastic Network Model Implementation

Let a graph *G* represent individuals (nodes) and their interactions (edges). Each individual (node) *i* is in a state *X*_*i*_*ϵ* {*S, E, I*_*pre*_, *I*_*sym*_, *I*_*asym*_, *H, R, F*}. The nodes adjacent (connected by a single edge) to an individual *i* defines its set of “close contacts” *C*_*G*_*(i)*, representing individuals with whom there is a regular interaction, such as household members, co-workers, friends, etc. This set cardinality stands for the degree of the network. At any time, individuals make either random contacts in their set of close contacts with probability *(1-p)*, or contact with individuals randomly sampled from the population at large, with probability *p*. These “global contacts” are one-off encounters with rare acquaintances or unknown individuals (for example at the supermarket, on public transportation means, at an event, etc.). Probability *p* defines the “locality” of an individual’s contacts’ network: for *p=0* an individual only interacts with close contacts, while *p=1* represents a uniformly mixed population. Transmission between an individual and his/her global contacts is referred to as global transmission, and transmission between an individual and his/her close contacts is referred to as local transmission. Finally, it is of great interest to consider the effect of ongoing introductions of the disease from outside the population. For example, an individual may interact with an infectious tourist, therefore introducing a new transmission chain.

### Non-Pharmaceutical Interventions (NPIs)

Social distancing and related NPIs (lockdowns, stay-at-home orders, “cocooning” of particular demographics, etc.) can be implemented by specifying a contact network. Dynamic scenarios can be explored by changing the structure of the contact network during a simulation. All model parameters are assigned to each node on an individual basis (arbitrary parameter heterogeneity). Varying the parameters as well as setting network properties such as the mean number of adjacent nodes (“close contacts”), allows modelling the degree of NPIs. Social distancing interventions may increase the locality of the network (i.e., decrease *p*) and/or decrease local connectivity of the network (i.e., decrease its degree).

### Model Equations

Let **1**_*Xi = Z*_ be an indicator function, equal to 1 if the state of node *i* is *Zϵ* {*S, E, I*_*pre*_, *I*_*sym*_, *I*_*asym*_, *H, R, F*}; equal to 0 otherwise. Then the propensities of state transitions (i.e. expected time to transition) for node *i* in Phase-1 model are given by the following equations:

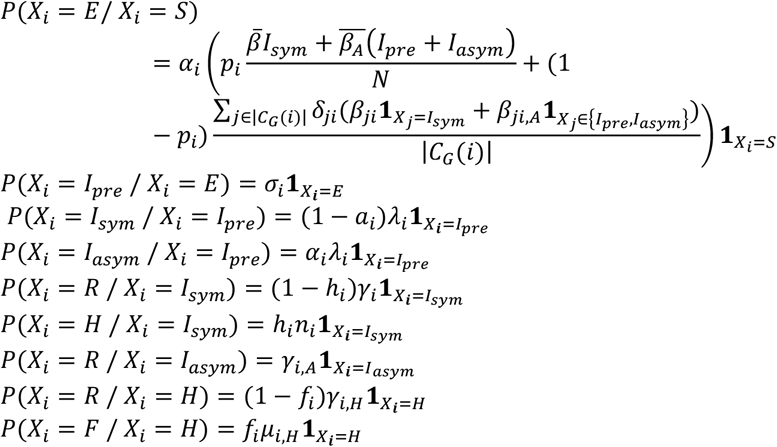

The propensities of state transitions for node *i* in Phase-2 model are given by the following equations:

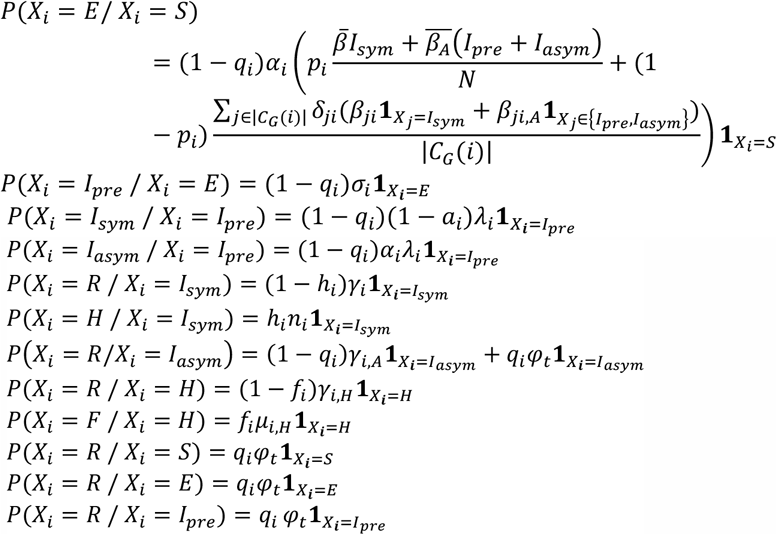

### Assumptions

- Regarding global interactions, every node in the population is equally likely to come into contact with every other node, and the population can be considered well-mixed. Thus, the contribution of the symptomatic subpopulation to an individual’s propensity for exposure is expressed through the mean transmissibility of the symptomatic individuals 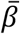 and the prevalence of symptomatic individuals in the overall population *I*_*sym*_*/N*. The contribution of pre- and asymptomatic infectious individuals involves the mean transmissibility of asymptomatic individuals 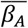 and their prevalence *(I*_*pre*_ *+ I*_*asym*_*)/N* ^5^.
- Regarding local transmission, transmissibility is considered on a pairwise basis, i.e., every directed edge of the contact network representing transmission from infected node *j* to susceptible node *i* is assigned a transmissibility *β*_*ji*_. The transmissibility of such an interaction is assumed in the proposed model to be equal to the transmissibility of the infected individual (i.e., *β*_*ji*_ *= β*_*j*_). The transition rate for any susceptible individual to become exposed due to local transmission is the product of that individual’s susceptibility and the total transmissibility of his/her infectious close contacts, divided by the size of his/her local network. Factor *δ*_*ji*_ appears in the calculation of the propensity for exposure due to local transmission and it is used to weight the transmissibility of interactions according to the connectivity of the interacting individuals, for example yielding a higher importance to a transmission between highly connected nodes-”superspreaders” (individuals who have more contacts also have more intense interactions). In this paper *δ*_*ji*_ is defined as

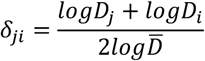

where *D*_*j*_ and *D*_*i*_ are the degrees of nodes *j* and *i*, respectively, and 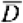 is the mean degree of the network. Similarly to the global transmission, the contributions of symptomatic and pre-symptomatic infectious contacts to the propensity for exposure are calculated separately^6^.
- The rate of re-susceptibility is assumed to be negligible (*ξ* is assumed to be equal to zero).
- The percentage *q* of the individuals that will be willing to participate in the mass vaccination and at the same time the vaccine will be effective on them is considered constant for all states.
- The number of vaccine-related deaths is negligible. This assumption is based on the fact that individuals contraindicated for vaccination have been totally excluded from the vaccination queue by a medical pre-screening.

## 4. Experimental Results

### Covid-19 background in Greece

In Greece, the first COVID-19 case was reported on February 26, 2020 and, soon after, numerous NPIs were implemented. A first nationwide lockdown took place from March 23 to May 4^th^, 2020 and a second, less stringent, was imposed on November 7^th^, 2020 and as of December 31^st^, 2020 is still ongoing. A timeline of the major NPIs imposed in Greece is illustrated in Table 1.

**Table 1.** Major NPIs implemented in Greece during the COVID-19 pandemic.

| Major NPIs | Starting Dates |
| --- | --- |
| School and University closures | 11/03/2020 |
| Leisure closure | 13-14/03/2020 |
| Private enterprises closure | 18/03/2020 |
| 1st national lockdown | 23/03/2020 |
| Masking recommendations (“Light masking”) | 04/05/2020 |
| Leisure – mass gathering containment | 24/08/2020 |
| Leisure closure | 29/09/2020 |
| Mandatory masking (“Heavy masking”) | 24/10/2020 |
| 2<sup>nd</sup> national lockdown | 07/11/2020 |
| Nursery schools – kindergarten – primary schools closure | 14/11/2020 |

The number of daily confirmed cases by date of sampling for laboratory testing, the more informative weekly new cases to tests percentage ratio (“positivity”) and the daily number of deaths as of December 31^st^, 2021 are depicted in Figures 3, 4 and 5, respectively. By December 31st, 2020, there were 138,850 diagnosed cases and 4,838 deaths. As of December 31^st^, 2021, the corresponding naive case fatality rate in Greece is 3.48% and the mortality rate is approximately 45 deaths per 100,000 population, ranking the country 126^th^ out of 171 worldwide and 17^th^ out of 50 Europe-wide^7^.

**Figure 3.**
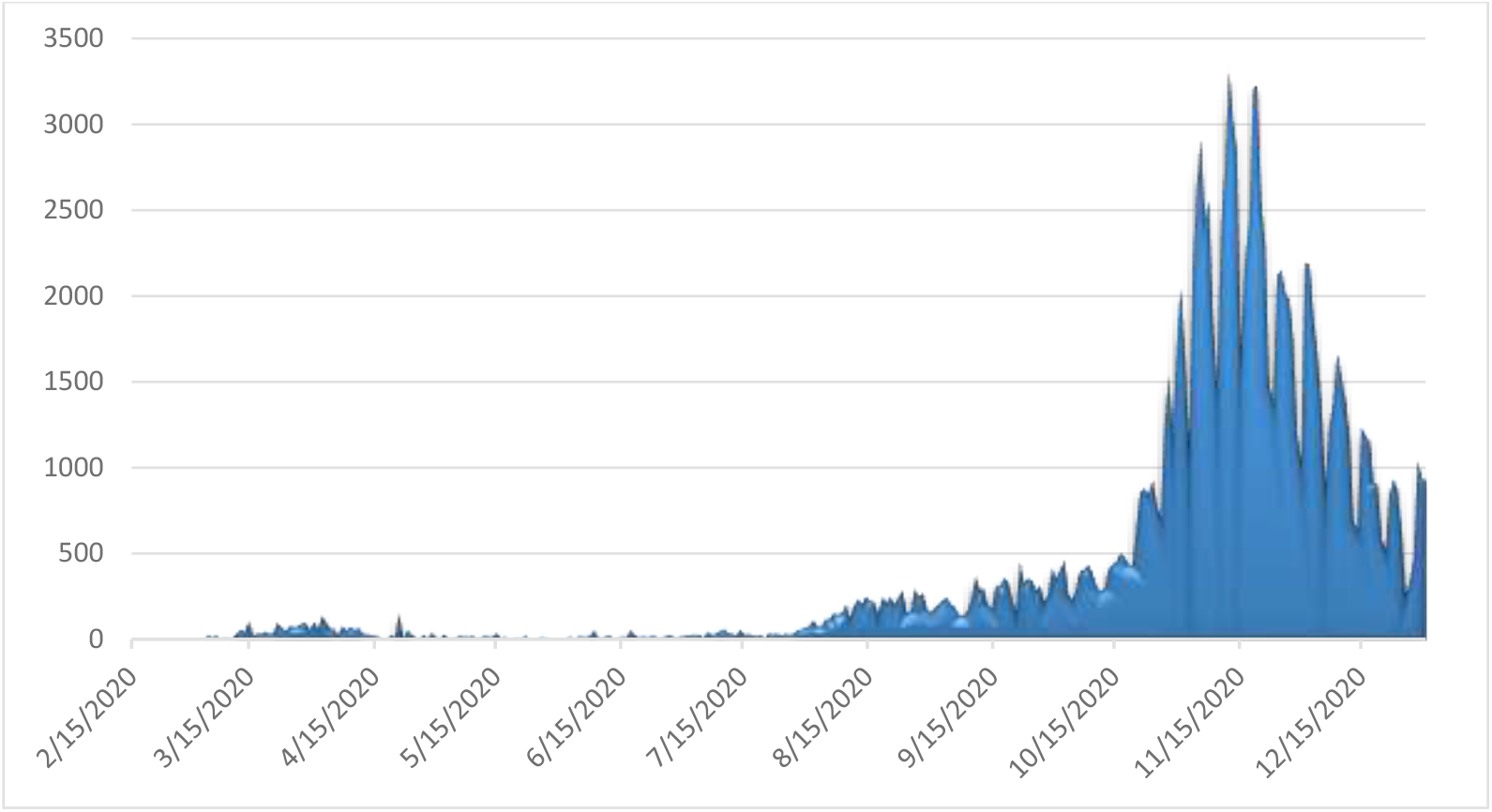
Daily number of laboratory-confirmed COVID-19 cases in Greece (Data available from ECDC^8^)

**Figure 4.**
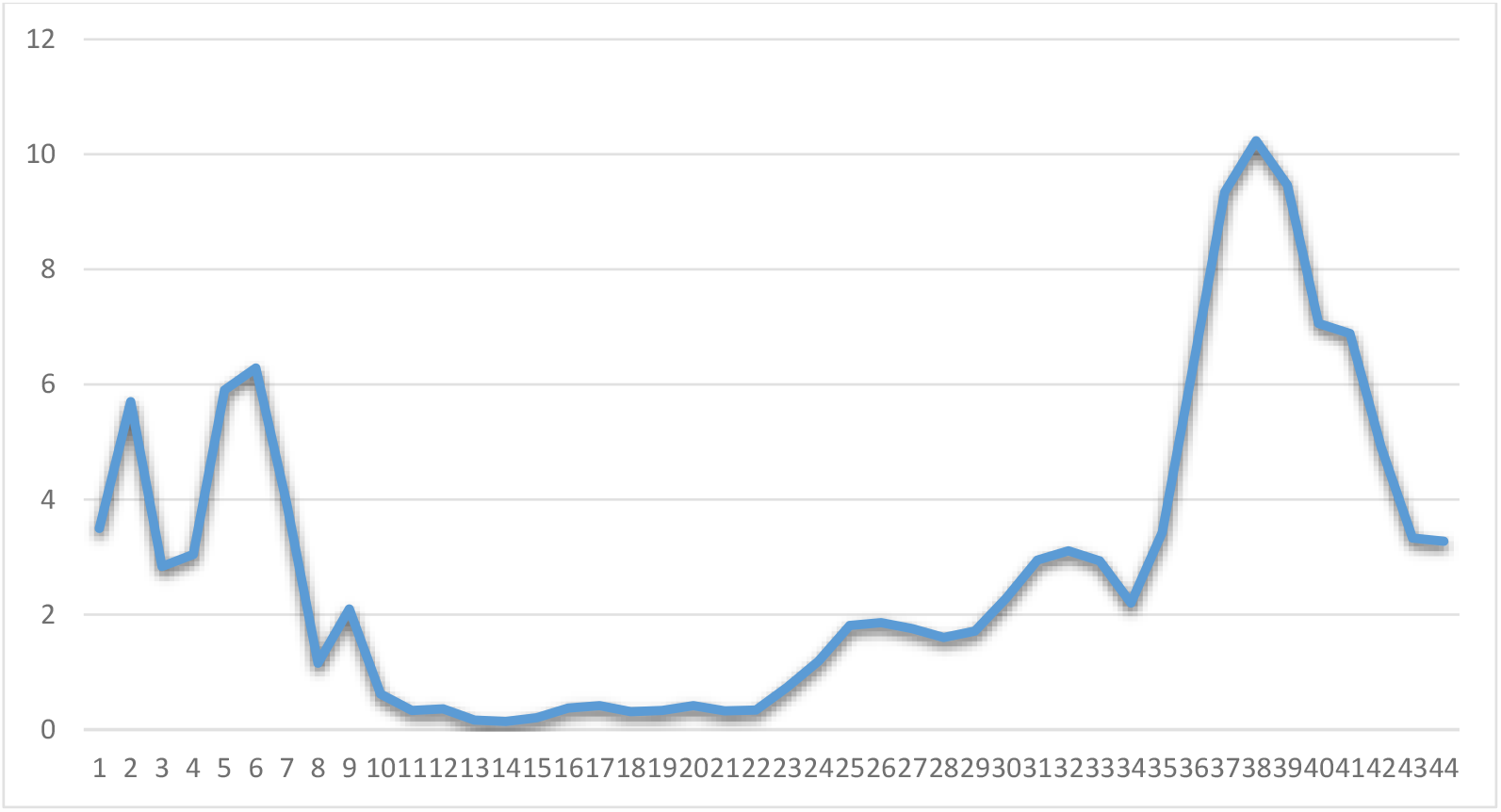
Weekly ratio of laboratory-confirmed COVID-19 new cases to tests in Greece (Data available from ECDC^9^)

**Figure 5.**
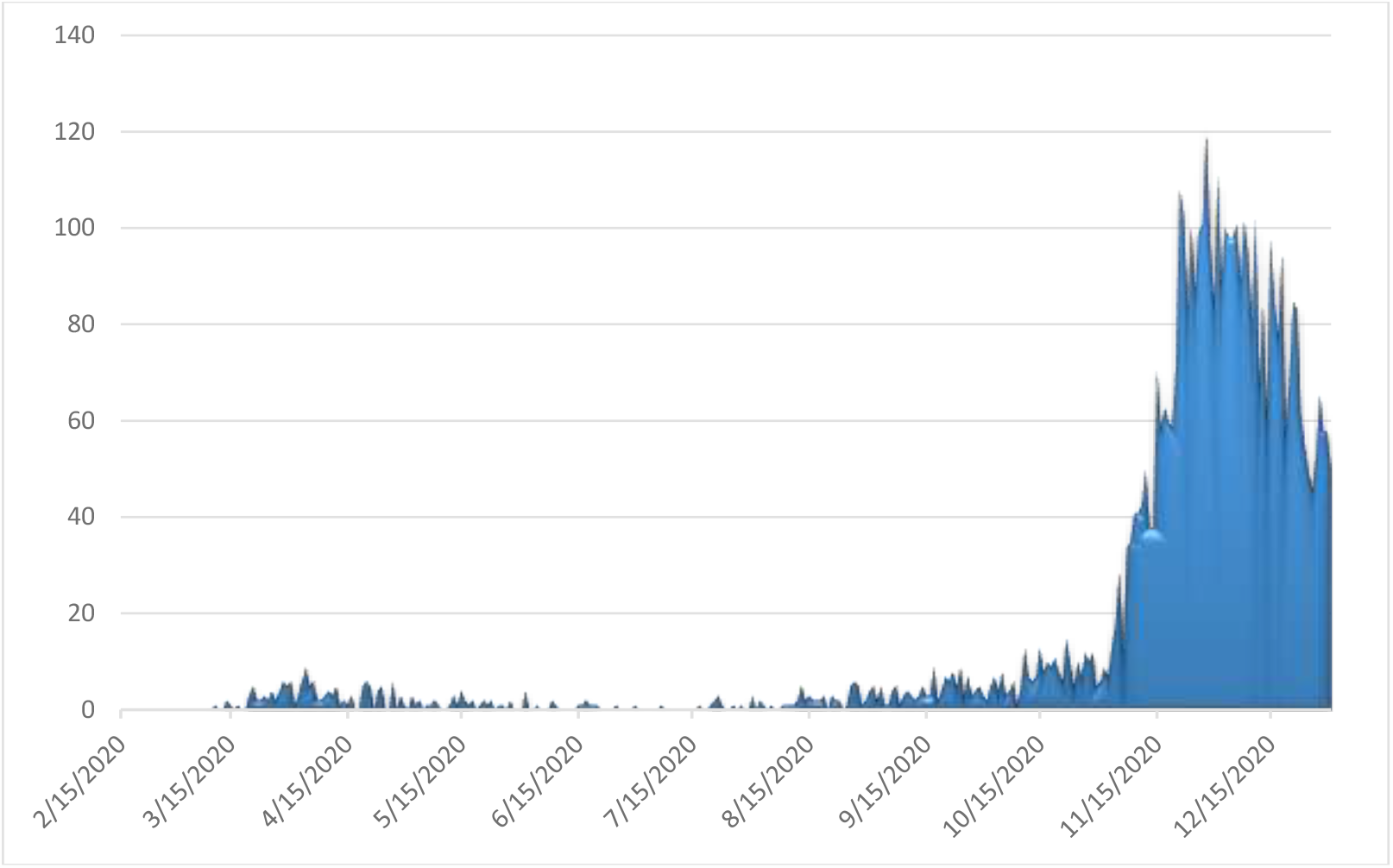
Observed daily confirmed number of deaths from COVID-19 complications in Greece (Data available from ECDC^10^)

### Scenarios

First, the approximately 10,800,000 individuals in Greece were decade-wise categorised into nine age groups, from “0-9 year” to “80+ years”, due to the strong age-dependence of COVID-19 progression and symptoms (lower attack rates and susceptibility among younger individuals and especially children [22]) and on the age-dependence of the contacts’ network structure (younger individuals may have 3-4 times more “global contacts” than older individuals). The size and relative frequency of each age group is based upon the most recent official national census data from the Hellenic Statistical Authority and are illustrated in Table 2. Given that the COVID-19 pandemic started in Greece less than a year ago, it is assumed that no changes in the population have occurred due to births, deaths, migration, etc.

**Table 2.**
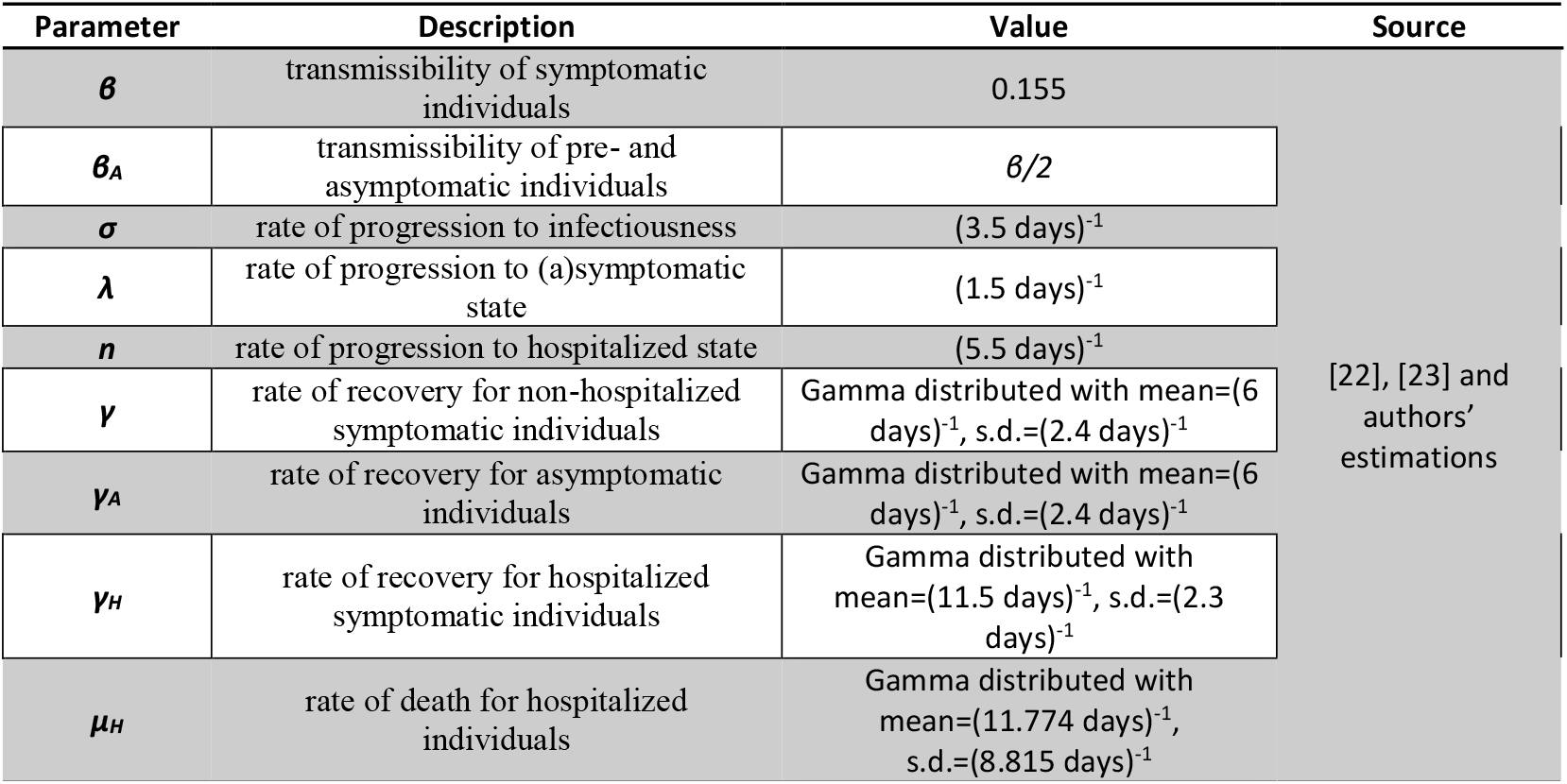
Models’ non age-dependent parameters’ values.

The outbreak of SARS-CoV-2 in Greece was simulated using Phase-1 SEIR model from the initiation of transmission until February 15^th^, 2021 and Phase-2 SVEIR model from February 15^th^, 2021 to June 30^th^, 2021. Transmission initiated on February 15^th^ 2020 as the earliest reported date of symptom onset among infected cases was February 20^th^ 2020, assuming that infection occurs approximately 5.5 days before (mean incubation period [22]). February 15^th^ 2021 was selected because the time period examined is exactly one year from the first recorded transmission, while it corresponds to the date by which the Greek government is planning to have achieved immunization through vaccination of approximately 1% of the total population^11^. It should be noted that the government plans to complete the mass vaccination of the population by June 30^th^, 2021.

The Greek government allowed retail partial opening (“click away” mode) in December 14^th^, 2020. Under this framework, three scenarios are assessed using Phase-1 model until February 15^th^, 2021:

- The *baseline* scenario, where the national lockdown and all NPIs are lifted on January 8^th^, 2021,
- A *“semi-lockdown”* scenario with school opening on January 8^th^, 2021, partial retail sector operating, universal mask wearing and teleworking,
- A “*rolling lockdown*” scenario with partial measures’ lift on January 8^th^ and a third imposed nationwide lockdown in February 2021 (“rolling” lockdowns policy).

The models’ parameter values estimates (either considered non age-dependent or adjusted to account for the non-homogeneous stratified age-groups distribution of the Greek population) are listed in Tables 2 and 3, respectively.

**Table 3.** Models’ age dependent parameters’ values.

| Age group | Group size<sup>12</sup> | Relative frequencies (%) | Relative susceptibility $\alpha$ [24] | % Infected asymptomatic $a$ [24], [25] | % Hospitalization $h$ [24], [26] and authors' estimations | %CFR for hospitalized $f$ [24], [26] and authors' estimations | Masking adherence (%) [22], [27] | Social distancing adherence (%) [22], [27] | Mean degree [23], [28] | $p$ [29] and authors' estimations |
| --- | --- | --- | --- | --- | --- | --- | --- | --- | --- | --- |
| 0 to 9 | 1,049,839 | 9.706 | 0.35 | 0.456 | 0.000011 | 0.0165 | 5-30 | 5-30 | 12.62 | 0.9 |
| 10 to 19 | 1,072,705 | 9.917 | 0.69 | 0.412 | 0.000114 | 0.0125 |  |  | 15.38 | 0.95 |
| 20-29 | 1,350,868 | 12.489 | 1.03 | 0.37 | 0.000495 | 0.025 |  |  | 12.89 | 0.7 |
| 30-39 | 1,635,304 | 15.119 | 1.03 | 0.332 | 0.003726 | 0.025 |  |  | 12.89 | 0.9 |
| 40-49 | 1,581,095 | 14.618 | 1.03 | 0.296 | 0.019272 | 0.0315 |  |  | 12.27 | 0.9 |
| 50-59 | 1,391,854 | 12.868 | 1.03 | 0.265 | 0.037107 | 0.08418 | 82.2 | 92.2 | 11.64 | 0.9 |
| 60-69 | 1,134,045 | 10.485 | 1.27 | 0.238 | 0.07067 | 0.1781 | 82.1 | 90.9 | 9.42 | 0.6 |
| 70-79 | 1,017,242 | 9.405 | 1.52 | 0.214 | 0.102833 | 0.38016 | 79.2 | 91.3 | 7.2 | 0.5 |
| 80+ | 583,334 | 5.393 | 1.52 | 0.192 | 0.131513 | 0.709 | 82.1 | 93.4 | 7.2 | 0.5 |

The major NPIs effects over time on the rates of infection in the population in the context of the three assessed scenarios are captured as follows:

- *Social distancing/teleworking/school closure*. Social distancing/teleworking/school closure effect is captured on one hand by the reduction of the individuals’ daily contacts (degree of the individual’s contact network) and on the other hand by the reduction of the locality parameter *p*. The graphs’ mean degrees and *p* by age group without any NPIs imposed are depicted in Table 3 and the decrease in their values by age group when social distancing/teleworking/school closure measures are imposed is illustrated in Table 4.
- *Lockdown*. Lockdown stringency is captured with a further decrease in the individuals’ contacts network degree and in the probability *p* of individuals coming into contact with those outside of their immediate network. This decrease differs by age group and the lockdown’s stringency and is illustrated in Table 4.
- *Mask wearing*. The factor by which *β* is reduced as a result of mask wearing depends on the utilization level (simple recommendation / strong recommendation / mandatory) as well as individuals’ adherence, which is age dependent. This decrease is illustrated in Table 4.

Phase-2 model’s initial states’ sizes are obtained from the optimal scenario in terms of fatalities yielded from Phase-1 model.

**Table 4.** Major NPIs effects on model parameters’ values.

| Major NPIs | Starting Dates | Network mean degree (age distribution) [22], [28]<br>and authors estimations | $\beta$ (age distribution) [23],<br>[27] and authors<br>estimations | $p$ (age distribution) [23], [29]<br>and authors estimations |
| --- | --- | --- | --- | --- |
| School and University closures | 11/3/2020 | 0-9: 6, 10-19: 12, 20-29: 11, 30-39:12, 40-49: 12, 50-59: 11, 60-69: 9, 70+: 7 |  | 0-19: 0.5, 20-39: 0.6, 40-69: 0.3, 70+: 0.2 |
| Leisure closure | 13-14/03/2020 | 0-9: 3, 10-19: 3, 20-29: 7, 30-39: 6 40-49: 6, 50-59: 6, 60-69: 8, 70+: 7 |  | 0-19: 0.3, 20-39: 0.4, 40-69: 0.3, 70+: 0.2 |
| Private enterprises | 18/03/2020 | 0-19:3, 20-29:6, 30-59: 5, 60-69: 6, 70+: 4 |  |  |
| 1st national lockdown | 23/03/2020 | 0-19: 3, 20-29: 5, 30-69: 3, 70+: 2 |  | 20-29: 0.05, Remainder: 0.02 |
| “Light” masking | 4/5/2020 | | 0-49: 0.95β, 50+: 0.8β | |
| Leisure – mass gathering containment | 24/08/2020 | 0-9: 12.62, 10-19: 15.38, 20-29: 12.89, 30-39:12.89, 40-49: 12.27, 50-59: 11.64, 60-69: 9.42, 70+: 7.2 |  |  |
| Leisure closure | 29/09/2020 | 0-9: 12.62, 10-19: 15.38, 20-29: 12, 30-39:11, 40-49: 11, 50-59: 11, 60-69: 8, 70+: 7 |  | 0-19: 0.6, 20-39: 0.7, 40-69: 0.5, 70+: 0.3 |
| «Heavy” masking +teleworking 50% | 24/10/2020 | 0-9: 13, 10-19: 16, 20-29: 12, 30-39: 10, 40-49: 10, 50-59: 10, 60+: 7 | 0-19: 0.5β, 20-50: 0.65β, 50+: 0.2β | 0-19: 0.5, 20-39: 0.6, 40-69: 0.4, 70+: 0.2 |
| 2<sup>nd</sup> national lockdown | 7/11/2020 | 0-9: 12.62, 10-19: 7, 20-29: 8, 30-69: 7, 70+: 3 |  | 0-9: 0.3, 10-19: 0.3, 20-29: 0.1, Remainder: 0.03 |
| Nursery schools – kindergarten – primary schools closure | 14/11/2020 | 0-19: 4, 20-29: 7, 30-69: 5, 70+: 3 |  | 20-29: 0.1, Remainder: 0.03 |

Regarding the mass vaccination strategy, as of December 31^st^, 2020 Greece had received two shipments of Pfizer/BioNTech Comirnaty of 93,600 vaccines in total. The plan is to receive from Pfizer/BioNTech, Moderna and AstraZeneca companies 919,250 vaccines until January 31^st^, 2021, 1,133,450 vaccines until February 28th, 2021, 2,995,800 vaccines until March 31^st^, 2021 and 2,200,000 vaccines until April 30^th^, 2021. Additional quantities, either from these three companies or from others that are developing their own vaccines, are scheduled to become available at the end of Spring 2021^13^.

The vaccination will take place in 1,018 centers (hospitals, health centers and other primary health care infrastructure) distributed all over Greece, while 65 mobile units will vaccinate individuals that cannot access vaccination centers (house-bound or institutionalized population). The vaccination design capacity is 5 individuals/hour/center, with centers operating 16h/day, 6 days/week. This is translated to 80 individuals vaccinated/center/day or 2,117,440 individuals/month nationwide. The vaccination prioritization the Greek government has announced places health workers and institutionalized elderly individuals first, followed by individuals aged over 65 and individuals with health problems, after whom comes the remainder of the population.

The available vaccine from Pfizer/BioNTech requires a reminder dose 21 days after the first dose, and its efficacy (after the reminder dose has been administered) is estimated to be around 90% for the individuals that are eligible for vaccination [30]. By February 15^th^, 2021, approximately 100,000 individuals are scheduled to be vaccinated, thus approximately 90,000 individuals will be immunized (moved to the recovered state). A 70% voluntary participation of the population is assumed based on statistical surveys conducted in Greece around mid-December 2020; with a 90% efficacy it is estimated that *q*=63%.

Then three scenarios with different vaccination rates are assessed:

- An optimistic scenario with φ_t_= 1/81,440 days (design vaccination capacity).
- A most likely scenario with φ_t_= 1/60,000 days
- A pessimistic scenario with φ_t_= 1/40,000 days

### Implementation

Phase-1 and Phase-2 models were built using the SEIRS+ modelling tool^14^, version 1.0.9. The code in Python is available on GitHub^15^. The size of the total population was set to 10,800 (a representative typical case, that is a factor of 1,000 from the population of approximately 10,800,000 [23]). One symptomatic node (1,000 cases) was seeded to the population at day 0 (February 15^th^, 2020). The 2-phase model yields results for 500 days (365 days for Phase-1 and 135 days for Phase-2 model) and the epidemic was further calibrated every 1,000 new infected cases over the first 9.5 months. One hundred runs were performed for each scenario for both phases.

Default interaction networks were used, constructed as FARZ graphs and processes. Stochastic network dynamics are simulated using the Gillespie algorithm, which is a common and rigorous method for simulating stochastic interaction dynamics [31]. Finally, ongoing exogenous introductions of the disease (imported new cases, e.g., tourists and land workers) are inserted to the population by manually interfacing with the model code during a run.

### Scenarios’ Results

Figure 6 shows the simulation results for a representative Phase-1 *baseline* scenario run. Replacing the nationwide lockdown with only recommendations for social distancing measures on January 8^th^, 2021 results in uncontrolled disease spread. The expected fatalities on February 15^th^, 2021 are 9,920 (95% CI = [8372, 11369]). It is obvious that this scenario is used only to prove that NPIs combined with vaccination are mandatory in order not to devastate the national health system.

**Figure 6.**
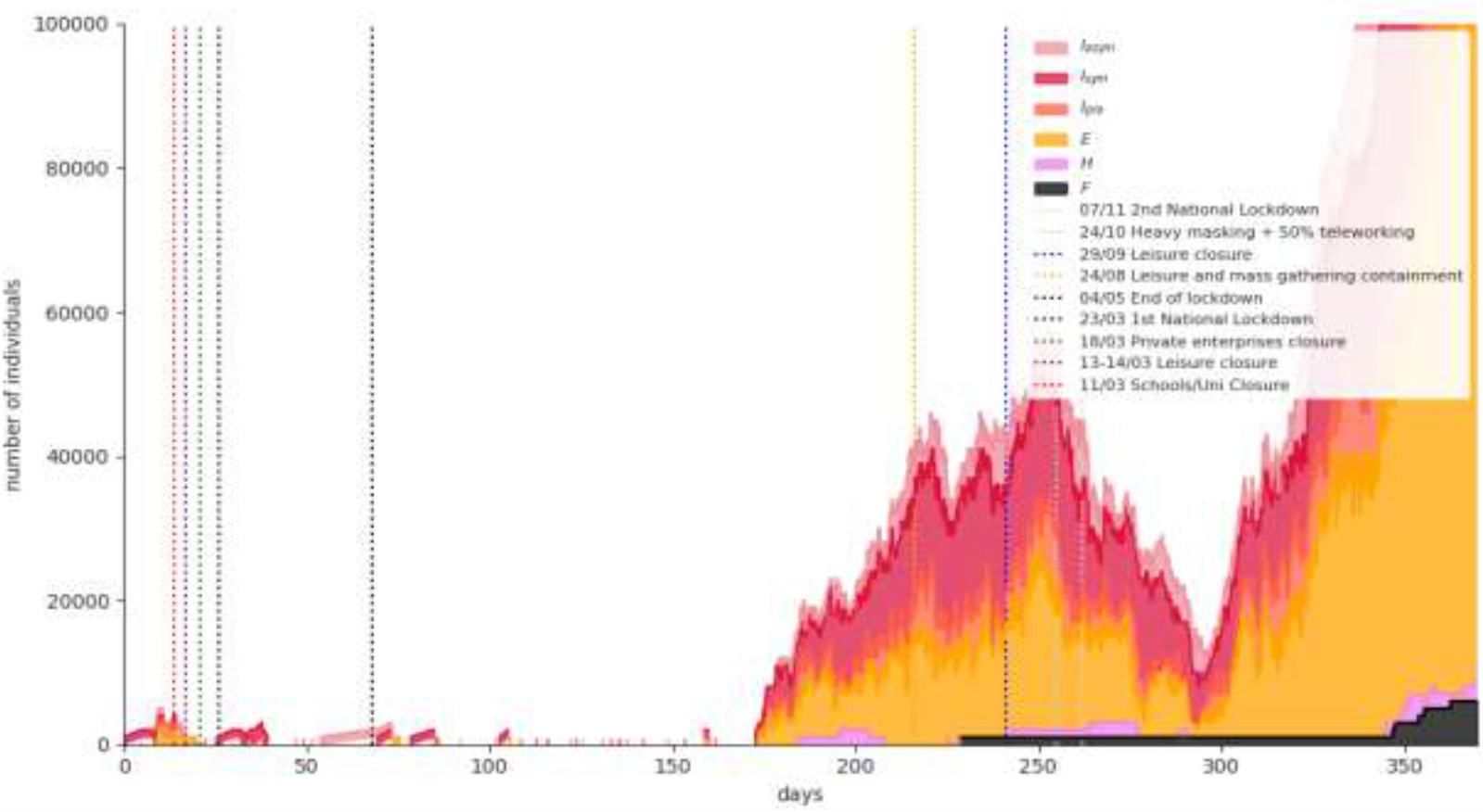
Simulation results for a representative Phase-1 *baseline* scenario run.

Figure 7 shows the simulation results for a representative Phase-1 *“semilockdown”* scenario run: universal masking and large-scale teleworking flattens the curve significantly, although an increase in the numbers of infected and hospitalized individuals is evident due to the population mobility caused by schools’ opening and retailing partial operation. The expected fatalities on February 15^th^, 2021 are 7,530 (95% CI=[5948, 9113]).

**Figure 7.**
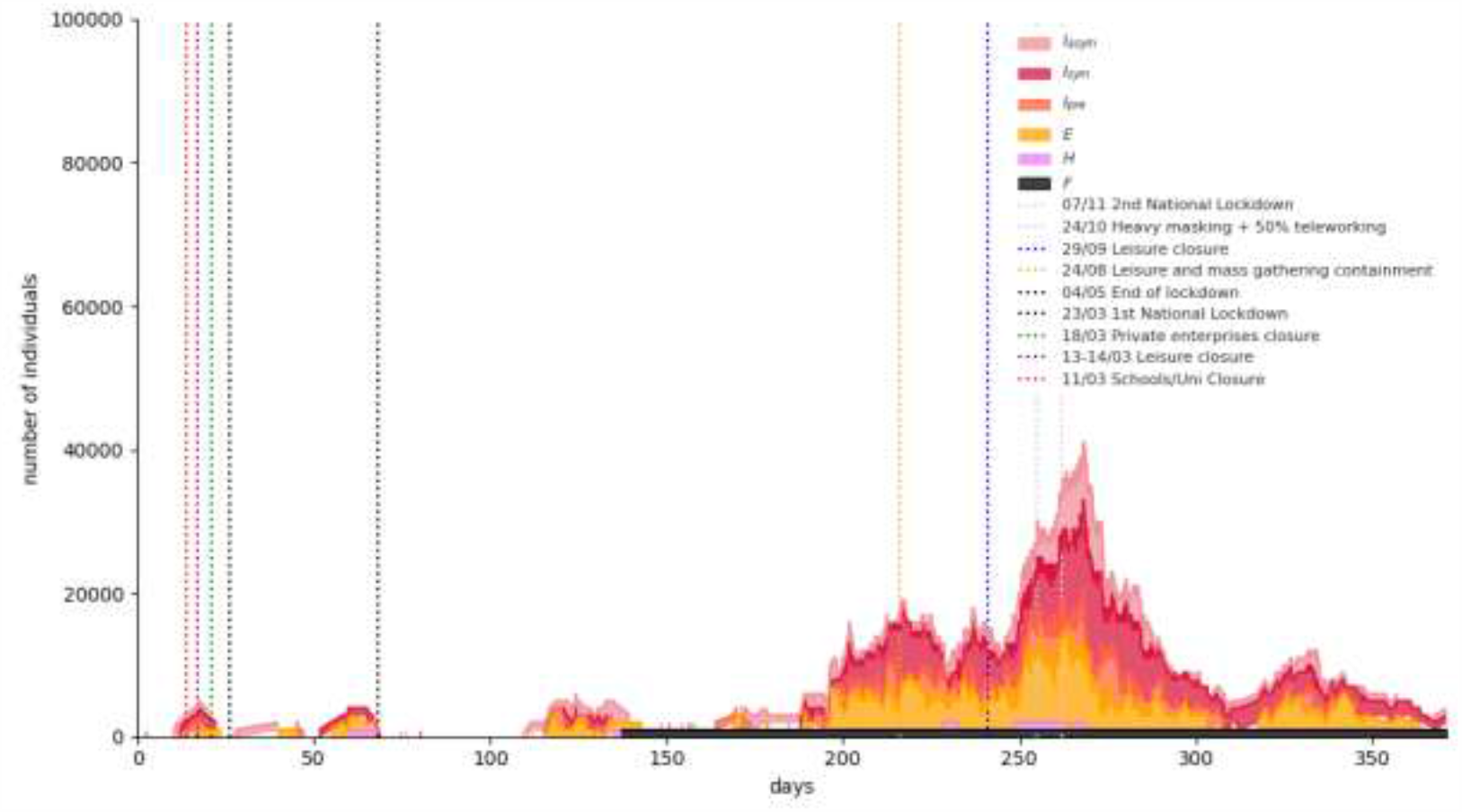
Simulation results for a representative Phase-1 “*semilockdown*” scenario run.

Figure 8 shows the simulation results for a representative Phase-1 “*rolling lockdown*” scenario run: A partial measures’ lift scenario at January 8^th^ with a third imposed nationwide lockdown in February 2021. The expected fatalities on February 15^th^, 2021 are 7,960 (95% CI=[6,350, 9,571]).

**Figure 8.**
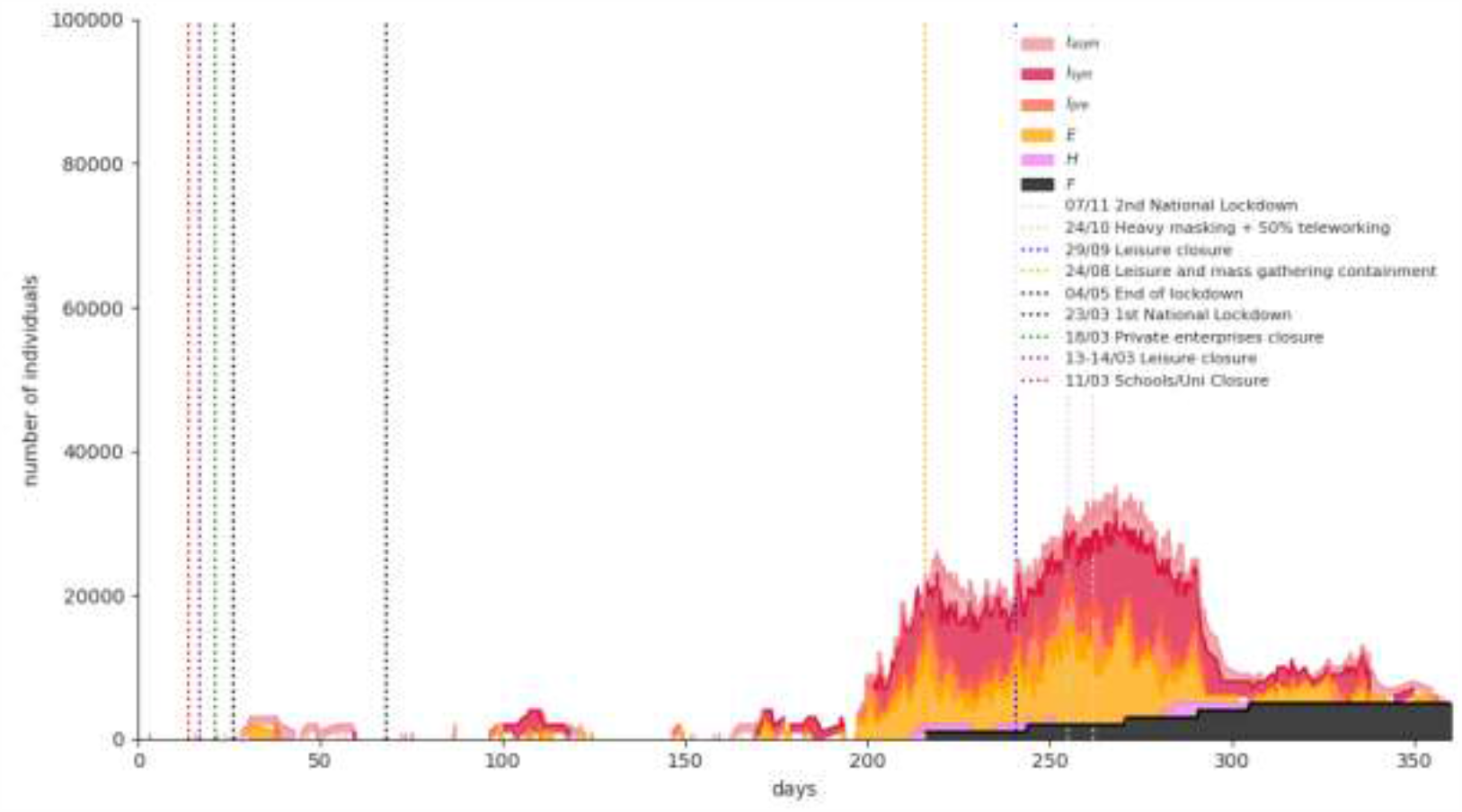
Simulation results for a representative Phase-1 “*rolling lockdown*” scenario run.

The results from the first phase indicate that the *“semi-lockdown”* scenario outperforms the *“rolling”* lockdown scenario (5.7% less expected fatalities), therefore Phase-2 model’s initial states’ sizes are obtained from it. More particularly, the worst *“semi-lockdown”* scenario run ending states’ sizes were selected as initial states’ sizes for the SVEIR model. Conditional on this selection, a gradual lift of NPIs is placed on April 1^st^, 2021.

The simulation results for a representative run for any of the three different vaccination rates are similar to the ones illustrated in Figure 9. The expected number of fatalities until June 30^th^, 2021 for the optimistic, most likely and pessimistic scenarios are 8,190 (95% CI=[8107, 8274]), 8,250 (95% CI=[8150, 8350]) and 8,290 (95% CI=[8200, 8380]), respectively, proving that the vaccines’ supply and vaccination rates are significant in minimizing fatalities from COVID-19, as it is the case in all epidemics [32].

**Figure 9.**
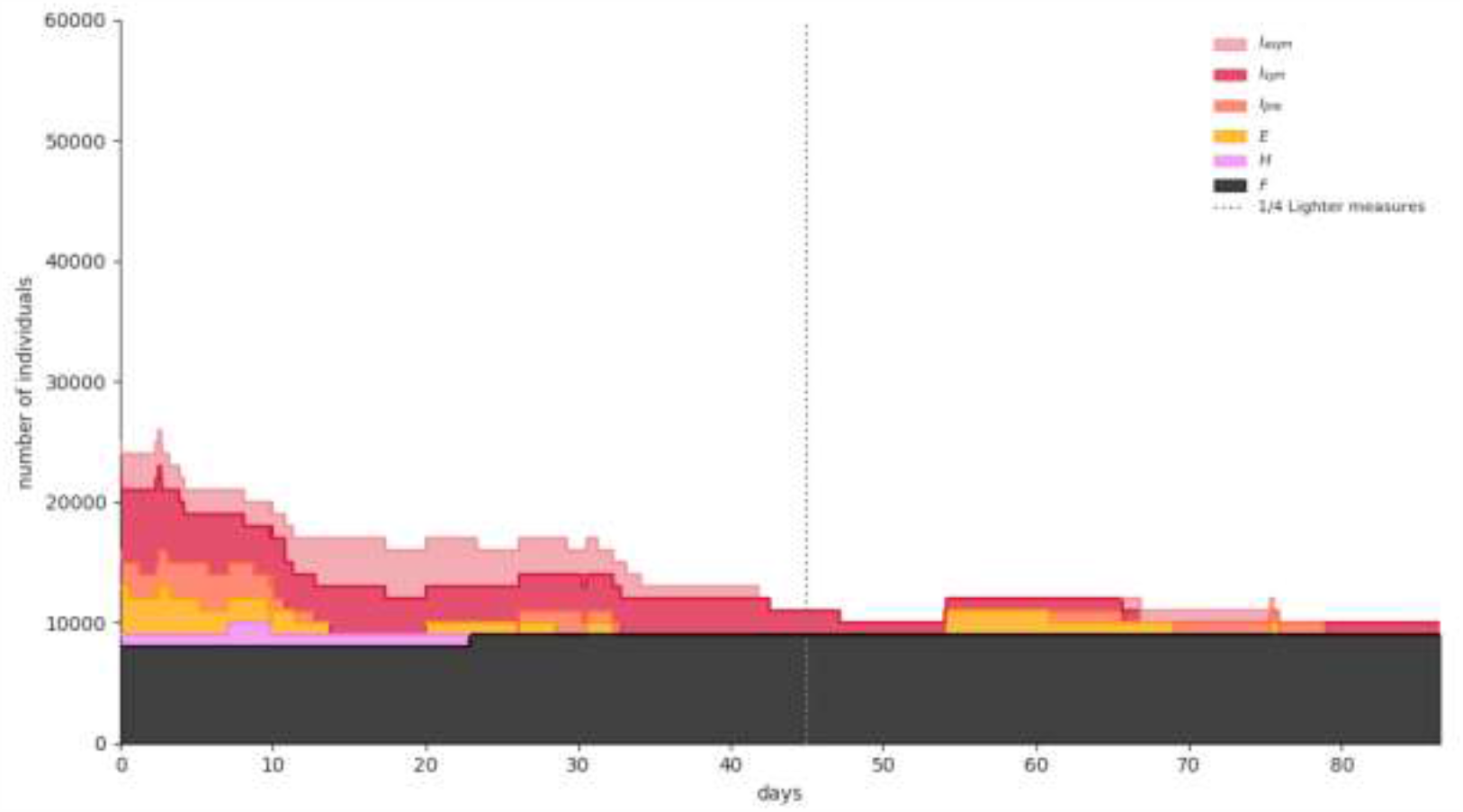
Simulation results for a representative Phase-2 scenario run.

## 5. Discussion

The 2-phase SEIR/SVEIR model proposed in this paper offers two major advantages in terms of COVID-19 simulation: First, since it is implemented on a stochastic dynamical network it mimics interactions between individuals in society more closely, rather than assuming uniform mixing as deterministic compartmental models do. Such an approach allows setting different model parameters for each node. Second, the NPIs and their introduction at different time periods are controlled in the simulation dynamically through the Python interface. This allows for policy interventions to be applied in response to possible changes in the spread of the epidemic (e.g., more stringent policies such as a local or national lockdown can be imposed when the number of new infections/ prevalence is above a threshold).

The Phase-1 model suggests a substantial impact of universal masking and large-scale teleworking, i.e., an adequate containment of the number of new infections, with better results in terms of fatalities than a “rolling” lockdown policy. A third stringent lockdown in February 2021 in Greece will eventually result in bringing the disease under control. However, its economic and social impacts will be enormous, not to mention the restrictions to individual freedoms. The two latter facts strongly support adopting an alternative solution, even with restrictions such as leisure opening not before April, 2021 and partial retail operating.

It is not easy to advocate an optimum strategy balancing efficacy with social and economic costs: a large number of minor NPIs that were applied in Greece from May 4^th^, 2020 until the beginning of September, 2020 did not have the results decision makers expected, considering the onset of the pandemic’s second wave. On the other hand, population fatigue during the second national lockdown imposed in November 2020 was one of the reasons that it proved less effective than the first imposed in March, 2020. Nevertheless, a mid-term decision support system remains indispensable in order to navigate a pathway to mass vaccination that will eventually contain COVID-19.

The analysis performed has several limitations. The lack of baseline pre-epidemic contact networks’ data implies that certain parameters’ values can only be guessed at. Even using SHARE COVID-19 July 2020 survey answers of the over 50 population, a recall bias might exist, albeit unclear as to direction. Additionally, the mass vaccination period raises many issues that are still not known: The vaccination prioritization is a very complex problem to solve. The vaccines that are gradually becoming available have different attributes, different administration schemes and different efficacies, varying from 62% to 94%. Finally, though a form must be filled by the individual before vaccination, this is unlikely to provide all the necessary information; vaccinators will be, for instance, to a large extent unaware of the susceptible/infectious status of all the asymptomatic individuals arriving at the vaccination system and it is certain that there will be a quantity of vaccines wasted per period.

## Data Availability

Publicly available data along with some preliminary first results of the SHARE COVID-19 survey conducted in Greece are used as input.

https://github.com/filipposfwt/UNIPI_COVID-19

## Acknowledgments

We would like to thank Professor Vana Sypsa, Department of Hygiene, Epidemiology and Medical Statistics, National and Kapodistrian University of Athens, Athens, Greece, for her valuable comments.

This work was supported by the European Commission under the Horizon 2020 Programme (H2020), as part of the project SHARE-COVID19 (Grant Agreement no. 101015924).

## Appendix: SHARE COVID-19 survey

The Survey of Health, Ageing and Retirement in Europe (SHARE) is a multidisciplinary and cross-national panel database of micro data on health, socio-economic status and social and family networks of about 140,000 individuals aged 50 or older (around 380,000 interviews). SHARE covers 27 European countries and Israel. As a reaction to the seriousness of the COVID-19 outbreak and the prolonged worldwide lockdowns, a special SHARE COVID-19 questionnaire was developed. This new questionnaire covers the most important life domains for the target population and asks specific questions about:

- Health and health behavior (General health before and after the COVID-19 outbreak, practice of safety measures, e.g. social distancing, wearing a mask)
- Mental health (Anxiety, depression, sleeping problems, and loneliness before and after the COVID-19 outbreak)
- Infections and healthcare (COVID-19 related symptoms, SARS-CoV-2 testing and hospitalization, forgone medical treatment, satisfaction with treatments)
- Changes in work and economic situation (Unemployment, business closures, working from home, changes in working hours and income, financial support)
- Social networks (Changes in personal contacts with family and friends, help given and received, personal care given and received).

In Greece, 3,901 SHARE’s panel respondents (57% females) were interviewed via a Computer Assisted Telephone Interview (CATI) from June 12^th^ to August 7^th^, 2020, answering the COVID-19 questionnaire. Sampling was performed by Hellenic Statistical Authority. Their geographical distribution in Greece’s 13 administrative districts (Figure A1) and their ages’ relative frequencies’ distribution are illustrated in Tables A1 and A2, respectively.

The COVID-19 questionnaire is available online^16^ [33]. Two questions were of particular interest for feeding the model, the one regarding wearing masks adherence and the one regarding social distancing adherence. The respondents’ answers are incorporated in Table 3.

**Figure A1:**
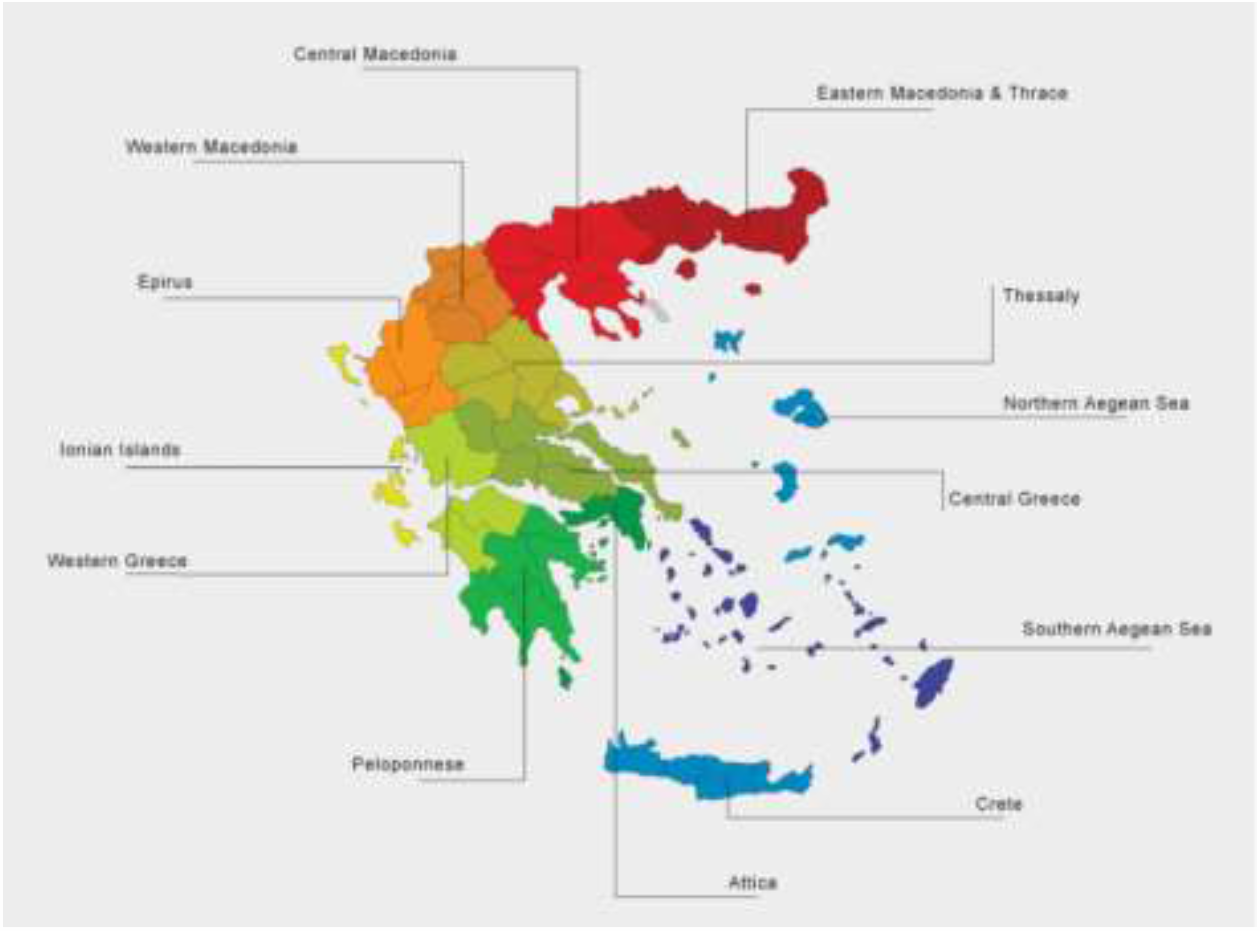
Map of Greece’s 13 administrative districts.

**Table S1:**
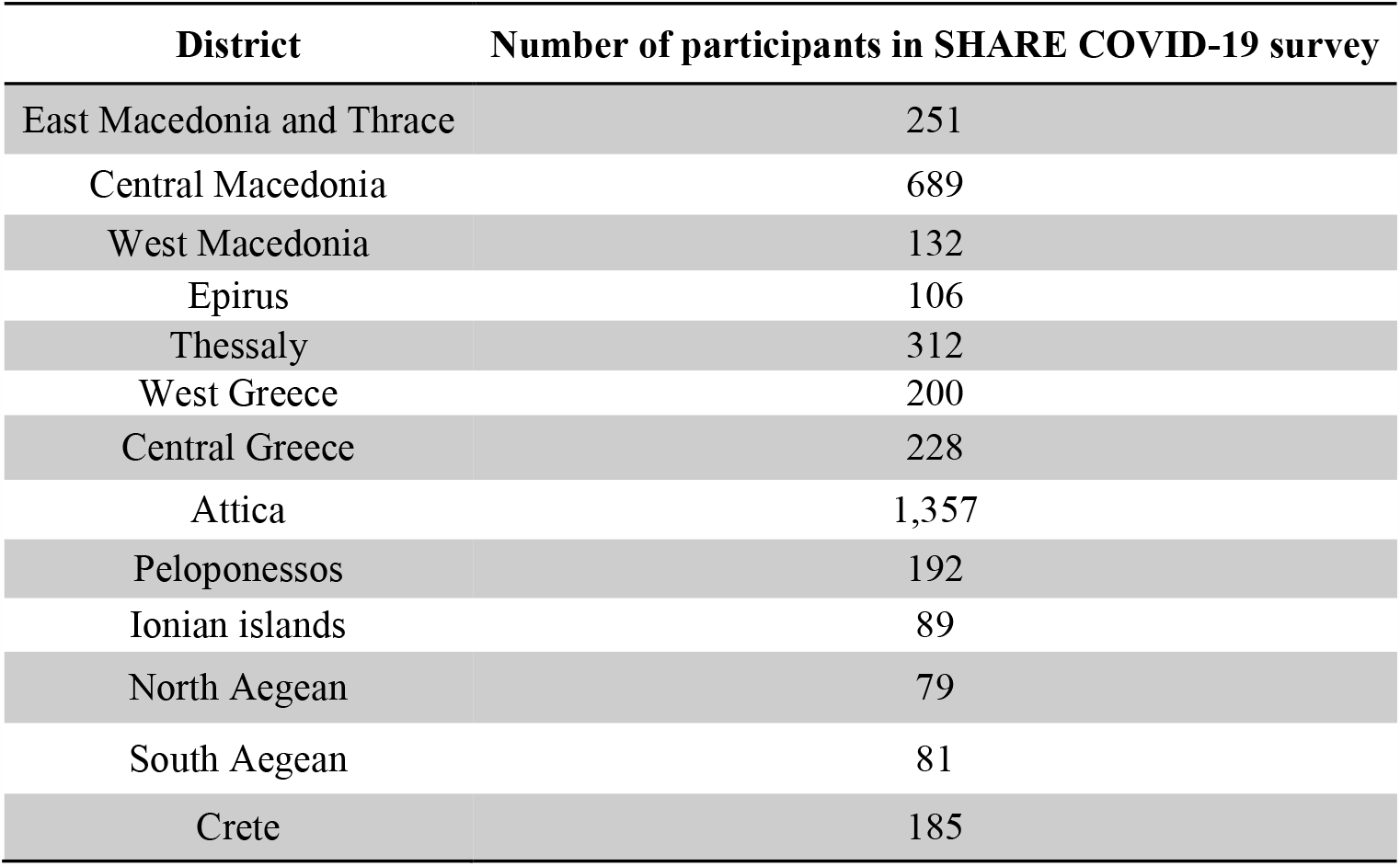
SHARE COVID-19 survey respondents’ geographical distribution.

**Table A2:** SHARE COVID-19 survey respondents’ age distribution.

| **Age** | **Relative Frequency** |
| --- | --- |
| 50-59 years old | 13.6% |
| 60-69 years old | 35.8% |
| 70-79 years old | 30.1% |
| More than 80 years old | 20.5% |

## Footnotes

1 https://covid19.who.int/?gclid=CjwKCAiArbv_BRA8EiwAYGs23Dd0_laqiTJ8UIMXAnWnWKCI9D_6gQlOsILFa_XMFoVPUPfnRkfxMhoCdi4QAvD_BwE

2 https://eody.gov.gr/epidimiologika-statistika-dedomena/ektheseis-covid-19/

3 https://github.com/ryansmcgee/seirsplus

4 https://github.com/ryansmcgee/seirsplus

5 https://github.com/ryansmcgee/seirsplus

6 https://github.com/ryansmcgee/seirsplus

7 Mortality Analyses - Johns Hopkins Coronavirus Resource Center (jhu.edu)

8 https://www.ecdc.europa.eu/en/publications-data/covid-19-testing

9 https://www.ecdc.europa.eu/en/publications-data/covid-19-testing

10 https://www.ecdc.europa.eu/en/publications-data/covid-19-testing

11 Ministry of Health.

12 Hellenic Statistical Authority (statistics.gr)

13 Ministry of Health.

14 https://github.com/ryansmcgee/seirsplus

15 https://github.com/filipposfwt/UNIPI_COVID-19

16 http://www.share-project.org/fileadmin/pdf_questionnaire_COVID-19/SHAREw8_COVID19_qnn_20200602_routing.pdf

